# Rare Variant Association Analysis Uncovers Involvement of *VNN2* in Stroke Outcome

**DOI:** 10.1101/2024.09.18.24313937

**Authors:** Estefanía Alcaide-Consuegra, Marina Mola-Caminal, Georgia Escaramís, Uxue Lazcano, Isabel Fernández Pérez, Joan Jimenez-Balado, Eva Giralt-Steinhauer, Elisa Cuadrado-Godia, Angel Ois, Ana Rodríguez-Campello, Marta Vallverdú, Aina Medina-Dols, Carmen Jiménez, Silvia Tur, Rosa M Díaz, Carlos David Bruque, Nuria Andreu-Somavilla, Irene González-Navarrete, Cristòfol Vives-Bauzà, Israel Fernández-Cadenas, Jordi Jiménez-Conde, Susanna Balcells, Raquel Rabionet

**Author notes:** Correspondence to Dr. Raquel Rabionet, Department of Genetics, Microbiology and Statistics, IBUB, IRSJD, Facultat de Biologia, Universitat de Barcelona, Barcelona, Catalunya 08028, Spain.

## Abstract

**BACKGROUND:** A stroke’s functional outcome presents vast variability among patients, which is influenced by age, sex, characteristics of the lesion, and genetic factors. However, there is very little knowledge about stroke recovery genetics. Recently, some GWAS (Genome-Wide Association Studies) have highlighted the involvement of common or low-frequency variants near or within *PATJ*, *PPP1R21, PTCH1, NTN4* and *TEK genes*, whereas the role of rare variants is still unclear. This study aims to identify the genetic contributions to differences in stroke outcome analyzing the effect of rare variants.

**METHODS:** We performed a pilot study analyzing 90 exomes of extreme good or bad recovery (modified Rankin Scale (mRS) at 3 months 0-1 vs 4-5) to select target genes involved in stroke recovery. To expand this study, 702 additional samples were sequenced by Targeted Next-Generation Sequencing capturing loci selected from the pilot study, GWAS studies and literature input. Here, we performed continuous (mRS 0-6) and dichotomous (mRS 0-1 vs 3-6) analyses, yielding one candidate gene. Protein structure and stability analysis were performed on selected variants.

**RESULTS:** Our work identified rare coding variants in *VNN2* associated with patients with a better stroke recovery (ΔDIC > 10, equivalent to p-value < 0.001). Six rare variants were predicted to significantly affect protein stability (ΔΔG > 1.6 kcal/mol), meanwhile, another variant, located in the active site, could affect the electrostatic surface.

**CONCLUSIONS:** *VNN2* could play a role in post-stroke inflammation altering the cell adhesion and migration of neutrophils during recovery.

## 1. Introduction

Worldwide, stroke is the second leading cause of death and adult disability. Approximately 1.1 million people in Europe suffer a stroke every year, and its incidence and prevalence are expected to increase along with the aging of the population^1^. The outcome of a stroke is affected by many factors including gender, age, stroke severity, size, and location. However, even adjusting for these factors, significant clinical heterogeneity remains. This heterogeneity can be partially explained by genetic variation affecting proteins involved in the stroke recovery process https://paperpile.com/c/bPJy0R/jEWX^2^.

Stroke recovery is a complex process that involves neuronal, vascular and immune responses^3^. Initially, after the ischemic event, in response to the injury, necrotic and dying neurons release damage-associated molecular patterns (DAMPs). These activate an innate immune response, including glial activation and infiltration of blood-borne immune cells into the brain. Microglia secrete matrix metallopeptidases (MMPs) that disrupt the integrity of the blood brain barrier (BBB), facilitating the invasion of macrophages and neutrophils. While this immune response is beneficial at stroke onset, if this pro-inflammatory response is extended, as it happens in aged individuals, it contributes to the participation of T-cells and a magnification of the immune response, and a worse stroke outcome^4^. Once this first response has taken place, poststroke recovery processes lead to restoration or compensation of function. These are based on the induction of key biological processes such as angiogenesis, neurogenesis, axonal sprouting, dendritic branching, synaptogenesis, and oligodendrogenesis^3^. An avenue to understand the relevance of all these processes is to identify key genes involved in stroke outcome, and the biological pathways that mediate the identified allele-outcome correlations.

Only a few studies have investigated the role of genome-wide genetic variation in ischemic stroke (IS) outcome, focusing on the common (>1%) genetic variation captured by classic GWAS (i.e. genotyping arrays)^5–8^. Studies looking at mid-term outcome (60 to 190 days) have identified two genome-wide significant associations, with variants near *PATJ*^5^, a gene related to tight junction formation and maintenance^9^, and variants regulating *PPP1R21*^6^, a gene involved in learning, memory and neuronal plasticity^10–12^. They also found suggestive association with *PTCH1*, *TEK*, and *NTN4*^6^. In addition, the analysis of the global effect of copy number variation, or genomic imbalance, on stroke outcome, yielded association of increased genomic imbalance with poorer stroke outcome^7^. Finally, another GWAS has uncovered associations of excitotoxicity related genes with short-term (24h) stroke outcome^8^, which is also correlated with 90-days outcome^13^. Meanwhile, the potential effect of rare variants, tackled by exome or whole genome sequencing, remains unexplored^2^.

Importantly, there are few to none widely accepted neuroprotective or neuroreparative drugs, nor personalized approaches to guide therapies to mitigate ischemic brain injury or enhance recovery^2^. Therefore, advances in the identification of novel outcome related variants and genes, coupled with the understanding of the pathways through which these genes affect stroke outcome, should provide critical knowledge for drug selection and allow for personalized therapy approaches.

The improvement of Next-Generation Sequencing (NGS) methods has allowed the study of both common and rare variants in complex diseases. Some studies have shown that rare variants could have higher impact on the structure, stability, or function of proteins than common variants^14,15^, explaining part of the heritability of complex traits^16^. However, association tests of individual rare variants require very large sample sizes, which are difficult to obtain for very specific phenotypes, such as a stroke’s functional outcome. To overcome this limitation, various rare variant association studies (RVAS) aggregate the effects of variants affecting the same biological entity (e.g. genes), and test for association of the aggregated variants with the phenotype. Other RVAS go further and consider more complex scenarios, such as heterogeneity of the variant’s effects (SKAT, SKAT-O), or variant-specific characteristics (BATI), providing increased statistical power^17–19^.

In the present analysis we explore the effect of rare variants on stroke outcome by applying BATI, which also integrates patients- and variant-specific characteristics as covariates. We performed a two-phase study, including exome sequencing in a small (n=90) but carefully curated cohort of extreme outcome phenotypes to select genomic regions that were further explored in a targeted sequencing analysis in 702 additional stroke cases. One gene, *VNN2*, harbored an excess of rare variants that potentially affect protein function in individuals with better outcome scores.

## 2. Methods

### Study Design

The study was divided into two phases: a first approach through a whole exome sequencing (WES) pilot study, and a follow-up by targeted resequencing analysis. The WES pilot study was performed on a selection of 90 cases matched by age, gender, and stroke type and location, divided into good (mRS 0-1) or poor (mRS 3-5) functional outcome. A second phase involved targeted resequencing analysis of 702 cases with a wider range of outcomes (mRS 0-6). Selected regions for resequencing included the top genes identified in the pilot WES study together with genes and regions around identified GWAS hits, as well as a few candidate genes from the literature (Supplementary bed file).

### Study Subjects

European ancestry patients with a diagnosis of IS according to World Health Organization criteria and available DNA samples were selected from the Spanish Stroke Genetics Consortium (GeneStroke) cohorts. The Pilot study included 69 patients from the BASICMAR study^20^ and 21 patients recruited at hospital Vall d’Hebron for the GRECOS^21^ and Geno-tPA^22^ studies. These patients were distributed in two subgroups based on their mRS scores at three months (0-1, “good outcome”, 49 patients vs 3-4-5 “poor outcome”, 41 patients), which were matched by age, gender, stroke location and type (Table 1). Inclusion and exclusion criteria are detailed in Supplementary methods. The targeted resequencing phase included 702 patients from the same cohorts (441 samples from BASICMAR study and 261 samples from GRECOS and Geno- tPA studies; Table 2 and Figure S1), fulfilling the same inclusion and exclusion criteria, except for the patients with unusual stroke (“other determined etiology” in TOAST). Exome sequencing data from the pilot study is deposited at the European Genome Archive (EGA dataset ID: EGAD00001004808). Targeted resequencing data is available upon request. The study was approved by the Institutional Review Boards of the participant hospitals (CEIm-PSMAR (2008/3083/I); IRB00002850 (PR(AG)157/2011)) and the CEIC of Fundació Sant Joan de Déu (C.I: PIC-82-17), and all participants provided written informed consent to participate: The research was conducted in accordance with the Helsinki declaration.

**Table 1.** Characteristics of the pilot phase samples.

| mRS (3 months) | Sex (female/male) | Age (range) | Age (mean) | LAA | CE | OD | Undetermined | Incomplete |
| --- | --- | --- | --- | --- | --- | --- | --- | --- |
| 0/1 | 24/25 | 53-87 | 73.26 | 12 | 22 | 0 | 13 | 2 |
| 3/4/5 | 20/21 | 51-90 | 74.4 | 11 | 20 | 0 | 6 | 3 |

LAA, large-artery atherosclerosis; CE, cardioembolism; OD, other determined; Undetermined, two or more causes; Incomplete, incomplete evaluation.

**Table 2.** Characteristics of the samples from the follow-up cohort.

| Characteristics | mRs at 3 months |  |  |  |  |  |  |
| --- | --- | --- | --- | --- | --- | --- | --- |
|  | 0 | 1 | 2 | 3 | 4 | 5 | 6 |
| Sex<br>(female/male) | 41/47 | 61/70 | 57/69 | 63/58 | 49/43 | 20/5 | 54/44 |
| Age (range) | 28-92 | 39-94 | 40-98 | 26-100 | 36-93 | 41-93 | 31-99 |
| Age (mean) | 70 | 72 | 74 | 76 | 77 | 77 | 81 |
| LAA | 13 | 23 | 29 | 23 | 18 | 4 | 9 |
| CE | 42 | 65 | 73 | 70 | 48 | 15 | 70 |
| OD | 18 | 31 | 19 | 17 | 11 | 2 | 8 |
| Undetermined | 3 | 4 | 2 | 5 | 9 | 1 | 10 |
| Incomplete | 10 | 7 | 3 | 6 | 6 | 2 | 1 |
| NA | 2 | 1 | 0 | 0 | 0 | 1 | 0 |
| Total | 88 | 131 | 126 | 121 | 92 | 25 | 98 |

LAA, large-artery atherosclerosis; CE, cardioembolism; OD, other determined; Undetermined, two or more causes; Incomplete, incomplete evaluation; NA, not available.

### Sequencing

For the pilot study, DNA obtained from peripheral blood lymphocytes was fragmented using a covaris system. Libraries were generated with TruSeqTM DNA Sample Preparation v2 Kits (Illumina, Inc., San Diego, CA, USA) followed by exome capture with NimbleGen SeqCap EZ Library SR v3.0 (Roche, Inc., Madison, WI, USA) in pools of 5 samples, which were sequenced to a 30-40x coverage on an Illumina HiSeq2500 at the Spanish national center for genomic analysis (CNAG).

For the targeted phase study, libraries were prepared from DNA from peripheral blood lymphocytes and captured with a custom agilent sureselect capture kit, using an in-house sample preparation system. Libraries were pooled and sequenced on Illumina Hiseq 2500 at the CRG-CNAG, to a targeted coverage of 150x.

### Bioinformatic Analysis

FASTQ raw sequences were processed following an in-house pipeline (Figure S2) including quality control with FASTQC, alignment with BWA-mem (http://bio-bwa.sourceforge.net/), duplicate marking with Picard (https://broadinstitute.github.io/picard/), samtools (http://www.htslib.org/) processing, local realignment base recalibration and variant calling with the Genome Analysis Toolkit (GATK; https://software.broadinstitute.org/gatk/). Variants were called by HaplotypeCaller using standard parameters except for min-pruning, which was set at 5 in the targeted analysis. Genome build 37 (GRCh37/hg19) was used as the reference genome. For exome sequencing, variants were further filtered with VQSR, following standard recommendations, while for the targeted resequencing, the resulting variants were hard-filtered following GATK recommendations (https://gatk.broadinstitute.org/) [QualbyDepth (QD) < 2.0; Quality (QUAL) < 30.0; StrandOddsRatio (SOR) > 3.0; FisherStrand (FS) > 60.0; RMSMappingQuality (MQ) < 40.0; MappingQualityRankSumTest (MQRankSum) < -12.5; ReadPosRankSumTest (ReadPosRankSum) < -8.0]. Multiallelic variants were filtered out with BCFtools (http://www.htslib.org/). The filtered VCF files were annotated with ANNOVAR (https://annovar.openbioinformatics.org/).

### Quality Control

In the pilot phase, 91 samples passed all quality criteria. In the targeted phase, three samples were removed based on inclusion and exclusion criteria, and another five were discarded because of missing information. Quality control on the pilot phase samples and the remaining 694 samples of the targeted phase was performed on the rvGWAS framework (available at https://github.com/hanasusak/rvGWAS). In both phases, variants with a genotyping call rate <95% were removed, as well as samples with >95% called variants, and outlier samples based on principal component analysis (PCA) and the number of called variants (Figure S3). This resulted in the removal of 13 samples in the targeted analysis, leaving a total of 681 samples for analysis. Finally, variants in top selected genes were manually curated by revision of variant calling files in IGV v. 2.8.0.

### rvGWAS framework

For downstream statistical analysis, only rare (European Allele Frequency < 0.01), nonsynonymous, probably damaging (CADD^23^ score > 20) exonic and splice-site variants were considered in both the pilot and the follow-up phase. In the pilot phase, association with the 3-month mRS was considered under a dichotomous model only (outcome mRS 3m 0-1 vs 3-5, analyzed with BATI and SKAT-O). In the follow-up, we considered two models, a continuous model (outcome mRS 3m 0-6, all 681 samples) and a dichotomous model (outcome mRS 3m 0-1, 219 samples, vs 3-6, 336 samples, totaling 555 samples). In both cases, analysis was performed with the Burden test, SKAT-O, KBAC, and BATI. Analyses were adjusted for age, sex, TOAST, and initial NIHSS. For the BATI test, variant effect (classified as loss-of-function or missense) was included as a covariate. An empirical significance threshold for the follow-up test (ΔDIC) was obtained performing 1000 simulations randomizing the 3-month mRS score. In each simulation, genes were ranked by their ΔDIC values, and the highest score was extracted, selecting the threshold at 0.1% significance level (ΔDIC_0.001_= 7.268). The default threshold for 0.1% of significance is ΔDICdefault > 10^19^.

### Protein Structure and Stability Analysis

Structural prediction for VNN2 was obtained from AlphaFold2 protein structure database prediction (European Molecular Biology Laboratory–European Bioinformatics Institute [EMBL-EBI], Hinxton, UK; https://www.alphafold.ebi.ac.uk/)^24^. The University of California, San Francisco (UCSF) Chimera program^25^ and the back-bone dependent rotamer library were used for structural interpretation and visualization^26^. Other tools used include TOPCONS^27^, PROSITE, InterPro and PFAM (see supplementary material).

The effect of variants on protein stability was calculated using FoldX (http://foldxsuite.crg.eu/)^28^. The structures were optimized to the FoldX force field command from the structure of VNN2 protein. The ΔΔG values were estimated as the difference between the energy of the wild-type and protein mutation (five replicates for each point variation). Values above 1,6 kcal/mol (twice the standard deviation) were considered to significantly destabilize the protein^29,30^.

## 3. Results

### Pilot study

The objective of the pilot study was to select approximately 100 candidate genes, potentially enriched in rare variants in either dependent or independent patients, to be followed up in a larger cohort by targeted resequencing. The final list of targeted regions which included regions selected based on the BATI and SKAT-O results and additional regions based on the literature, are provided in Table S1.

### Targeted NGS study highlights *VNN2* as a candidate gene

The analysis under the continuous outcome model did not yield any gene with significant enrichment of rare coding variants (Table S2 and S3). However, *TMPRSS7*, the top gene in the BATI analysis, showed a ΔDIC value of 6.9, close to the 1% empirical significance threshold (ΔDIC_empirical_ = 7.268).

In the analysis under the dichotomous model, we compared patients with a 3- month mRS of 0 or 1 with those with a 3-month mRS score over 3, using the following test and corresponding thresholds: Burden, SKAT-O, KBAC (p-value < 0.05), HBMR (Bayes Factor > 10) and BATI (ΔDIC_0.01_ < 7.268). Analysis with BATI showed one gene, *VNN2*, to be significantly enriched (ΔDIC*_VNN2_* < 10.87; ΔDIC_empirical_ = 7.268; ΔDIC_default_ = 10) for rare variants in patients with better outcome (Table 3). While not significant, *VNN2* was also among the top genes identified in all other tests (Tables S4, S5 and S6).

**Table 3.**
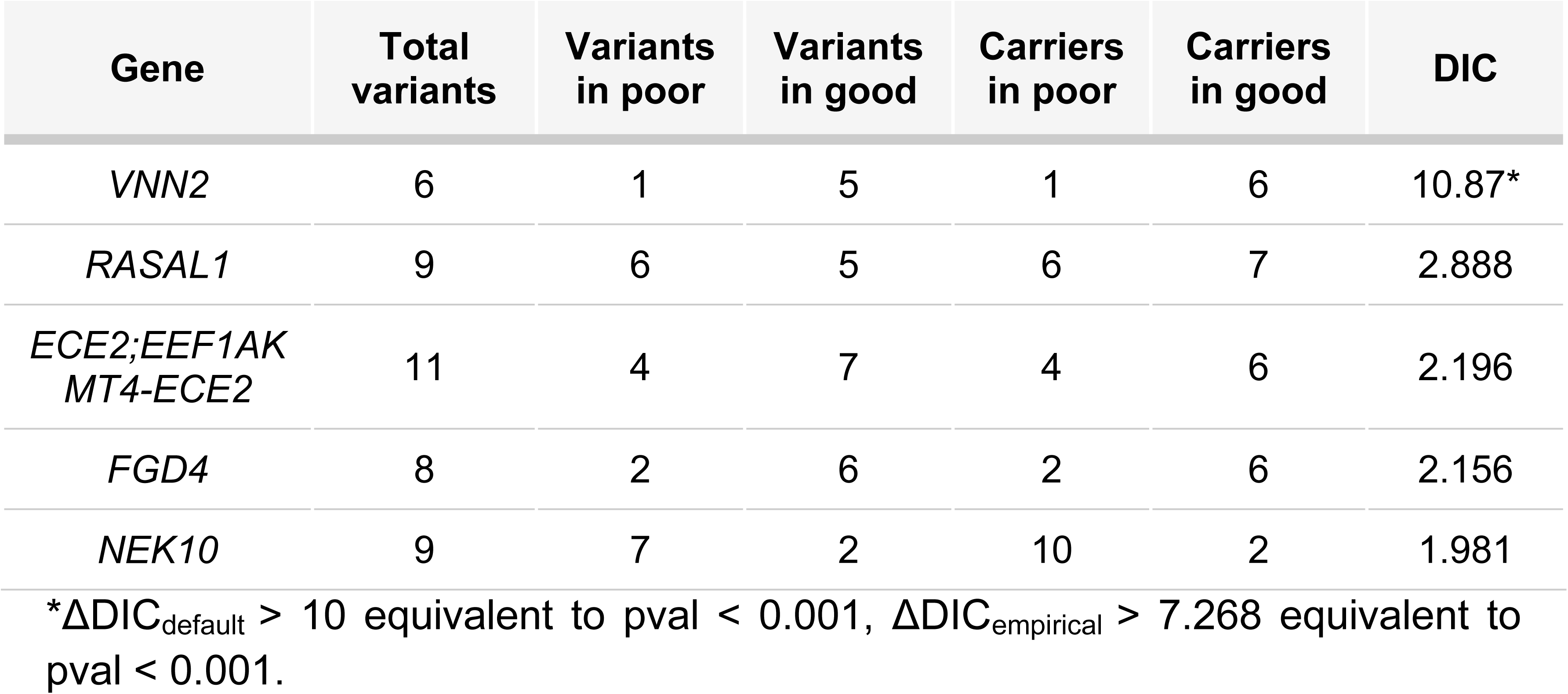
Top genes in the rare variant association study (dichotomous model) according to BATI. . The results were performed in a dichotomous model using mRs at 3 months 0-1 vs 3-6.

### *VNN2* variants identified in IS patients affect protein stability

Six *VNN2* variants were identified in the follow-up cohort, five of which were identified in six patients with good recovery, and one in one case with poor recovery. In addition, in the pilot study we had identified an additional two variants in cases with good recovery, while no variants were present in patients with poor recovery.

We explored the potential effect of the identified *VNN2* variants *in silico* (Table 4), and all of them were predicted to have a significant effect on the protein, either by affecting its stability, by altering its electrostatic surface or by truncating the protein.

**Table 4.** VNN2 protein evaluation. CADD 1.3. ^30^

| Protein Variation | Domain | Protein Effect | Protein Stability $\Delta\Delta\text{G}$ values | CADD (v 1.3) |
| --- | --- | --- | --- | --- |
| p.(S46F) | CN hydrolase | Stability | $1.99 \pm 0.09$ | 23.6 |
| p.(L53P) | CN hydrolase | Stability | $3.65 \pm 0.03$ | 25.8 |
| p.(V174M) | CN hydrolase | Stability | $1.7 \pm 0.27$ | 27 |
| p.(R205S) | CN hydrolase | Stability | $2.72 \pm 0.09$ | 22.7 |
| p.(A265T) | CN hydrolase | Stability | $5.9 \pm 0.09$ | 25 |
| p.(G284C) | CN hydrolase | Stability | $1.96 \pm 0.30$ | 23 |
| p.(R393Q) | Vanin C | Electrostatic Surface | $-0.18 \pm 0.06$ | 20.8 |
| p.(R393*) | Vanin C | NMD; GPI Anchor | - | 34 |

Six out of eight variants are located in the CN hydrolase domain (Figure 1A and 1B). Two variants, p.(Ser46Phe) and p.(Leu53Pro), are in the first alpha helix. Both substitutions are expected to have a clear structural impact on the helix. In one case, by replacing a small proline for a bulky, aliphatic leucine; in the other, a very large, aromatic, non-polar amino acid (Phe) for a tiny, polar one (Ser). This substitution (p.(Ser46Phe)) is predicted to create a steric effect that would compromise its interaction with the Aspartic residue in position 49 (Figure 1C). On the other hand, variants p.(Val174Met) and p.(Arg205Ser), located in the beta-pleated sheets A and B, are predicted to affect the interaction of their corresponding residues with the threonine residue at 198 and the phenylalanine at 199, respectively, destabilizing the region between these beta sheets (Figure 1D). Variant p.(Ala265Thr) is also predicted to cause a steric shift destabilizing the protein (Figure 1F), as it causes the substitution of a larger uncharged polar residue (Thr) for a small nonpolar aliphatic residue (Ala) in an internal hydrophobic region of the protein close to the active site. Finally, p.(Gly284Cys), the last variant in the CN-hydrolase domain, located at the end of the beta B fold sheet, would cause an effect on the adjacent loop affecting its mobility (Figure 1G).

**Figure 1.**
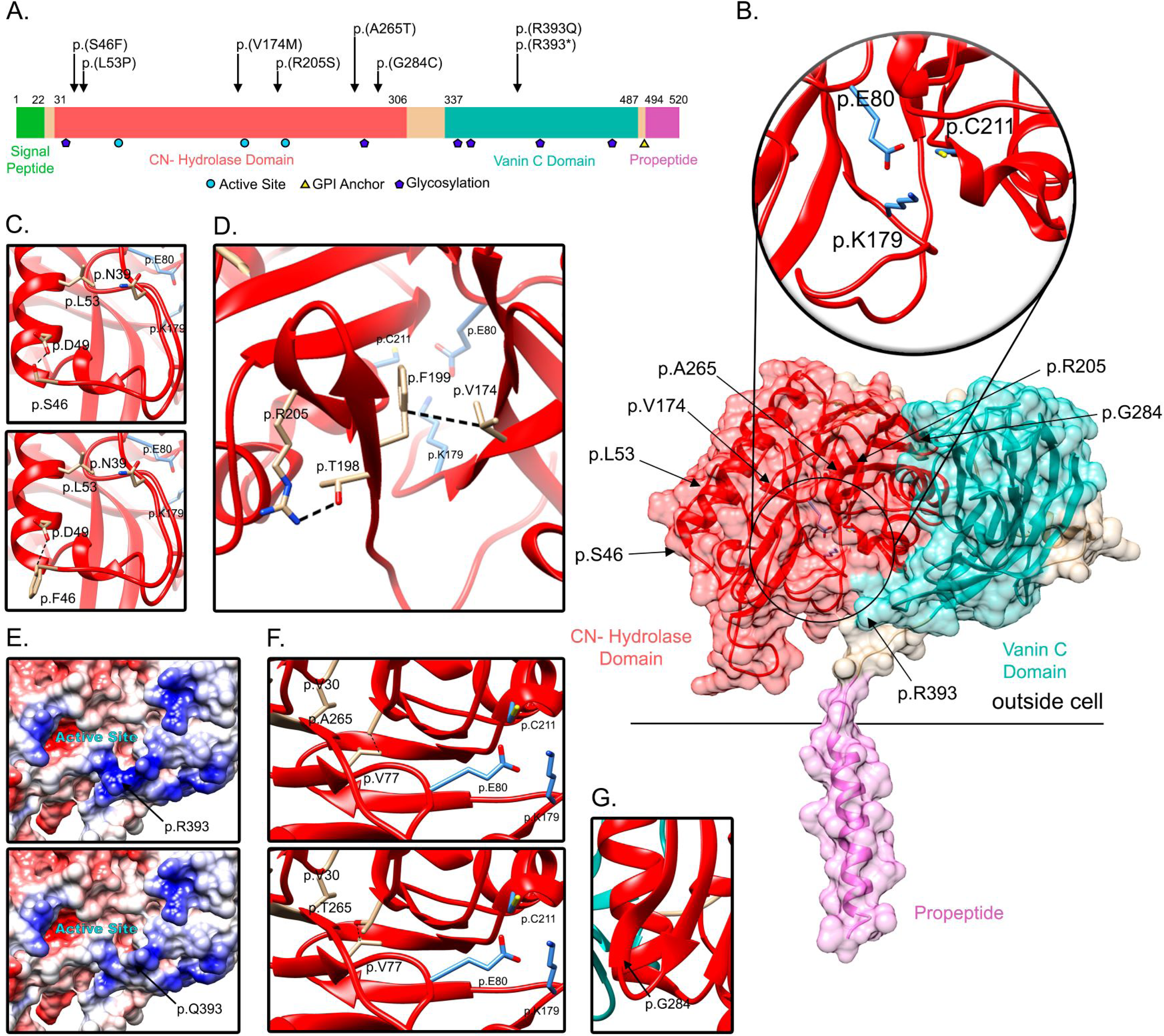
VNN2 protein structure and rare variants identified by WES and Targeted NGS analysis. (A) Lineal protein domains of VNN2. (B) Structural analysis of variants in VNN2 protein. Human protein structure prediction with AlphaFold2. In the upper panel, a zoom of the active site of VNN2 is shown. The lower panel shows the structure of VNN2 protein. In red, the CN Hidrolase Domain. In light blue, the Vanin C Domain. In pink, the propeptide/transmembrane region. Black arrows indicate the variants analyzed. (C, D, E, F, G) Zoomed regions highlighting conformational changes due to the amino acid variants.

The remaining two variants affect the same amino acid in the Vanin C domain. These variants are located in the exon 5 and may trigger nonsense-mediated decay (NMD). Even if NMD does not occur, the variant would still remove two thirds of this domain as well as the propeptide, including the loss of the GPI anchor. Moreover, the variant p.(Arg393Gln) is predicted to cause a change in the surface electrostatic charge of the protein. It is located at the access of the substrate to the active site, in a stretch of 4 consecutive arginines, conserved between species, that confer a positive charge to this region, which is affected by the substitution of an uncharged glutamine for the electropositive arginine (Figure 1E).

## 4. Discussion

After a stroke, the interaction of different environmental and genetic factors may define the functional outcome of the cerebrovascular accident. Although GWAS studies have successfully identified common variants involved in stroke recovery, rare variants have not been explored yet.

Here, for the first time, we performed an association study for rare variants involved in stroke recovery. This analysis involved a targeted analysis of 100 genomic regions in 702 patients, considering both a continuous (3-month mRS 0-6) and dichotomous (3-month mRS 0-1 vs 4-5) outcome variable model. Analysis was performed using SKAT-O and the BATI rare variant association test, which allows the integration of patient- and variant-specific features as covariates ^19^, and was proposed to have an improved power for the identification of the risk genes, especially in architectures with high genetic heterogeneity. BATI analysis yielded one gene, *VNN2,* significantly (ΔDIC > 10, equivalent to p-value < 0.001) associated with functional independence at 3 months.

*VNN2* (Vanin-2)^31^ encodes a glycosylphosphatidylinositol (GPI)-anchored extracellular protein also known as GPI-80^32^. VNN2 has been identified to play a role in the cellular adhesion and transmigration processes of human neutrophil extravasation^32^. While the precise role of VNN2 in this multi-step process remains to be defined, it seems to take place in the transition from rolling to firm adhesion^32–34^.

On the other hand, VNN2, together with VNN1 and VNN3, belong to the vanin protein family, characterized by their pantetheinase activity. Pantetheinase hydrolyzes pantetheine to pantothenic acid (vitamin B_5_) and cysteamine, activating the stress pathway and inflammation. While this activity is stronger in VNN1, VNN2 also has this capability through its CN-Hydrolase domain^35,36^.

We have identified eight rare variants in *VNN2*, seven of them present in patients with a good stroke outcome, while the remaining variant was present in a patient with poor outcome. *In silico* protein structure analysis predicted an effect in protein stability for the six missense variants located in the CN-Hydrolase domain (Table 4), indicating a potential effect of these variants through the modification of VNN2’s enzymatic activity. While the remaining missense variant, (p.(Arg393Gln)), is located in the vanin domain, and was not predicted to affect protein stability, it was predicted to affect the electrostatic surface charge, potentially limiting the access to the nearby active site. The remaining variant, p.(R393*), would lead to a truncated protein sequence lacking the propeptide sequence and the GPI anchor site. However, given the location of the nonsense mutation, it would be expected to lead to NMD and absence of protein. Therefore, all the heterozygous identified variants could lead to a reduction in VNN2 function, which could be associated with a lower inflammatory response.

The role of inflammation and immune mechanisms in the stroke recovery process is being increasingly recognized. Neutrophil extravasation and infiltration during inflammation are major contributors to poor ischemic outcomes^37^. Neutrophil interaction with endothelial adhesion molecules, a process in which VNN2 could be involved, together with the ischemic environment, is reported to shift the neutrophil phenotype from the protective N2 to more damaging N1 phenotype^37^.

It is possible that the rare VNN2 variants identified in these individuals could affect its function in the neutrophil extravasation process, or even the shift from N2 to N1 neutrophils and therefore reduce the infiltration of neutrophils into the brain parenchyma^37^. Alternatively, the variants could lead to a reduced pantetheinase activity, conferring higher resistance to oxidative stress and leading to reduced levels of inflammation.

To summarize, we have identified an excess of rare variants, predicted to affect protein stability or function, in stroke patients with a low mRs score at the 3- month evaluation, classified as a good functional outcome. We hypothesize that the loss or reduction of VNN2 activity could lead to lower inflammation levels. It has been shown that while this inflammation process is a necessary event, excessive inflammation can be detrimental in the recovery process^4^. A lower neutrophil extravasation or protection against oxidative stress caused by reduced VNN2 activity could lead to a reduced inflammatory response and allow for a better outcome.

In conclusion, we present the first study of rare variants involved in stroke recovery, highlighting a possible relationship between stroke outcome and rare variants in VNN2. This protein could act on oxidative stress response, or cell adhesion and migration of neutrophils contributing to a good outcome after stroke. Therefore, VNN2 might be a novel therapeutic target for stroke recovery. Nonetheless, to expand our knowledge of the rare variant architecture of IS recovery, a cohort amplification is required to improve the detection of rare variants, the identification of novel genes, as well as increasing captured regions with WES data. These insights would help to improve statistical power in both the continuous and dichotomous models.

## Data Availability

Exome sequencing data from the pilot study is deposited at the European Genome Archive (EGA dataset ID: EGAD00001004808). Targeted resequencing data is available upon request.

## Acknowledgments

We thank all the patients who suffered an ischemic stroke and allowed us to use their data in the study.

## Nonstandard abbreviations and Acronyms

- Damage-associated molecular patterns (DAMPs)

- Matrix metallopeptidases (MMPs)

- Blood brain barrier (BBB)

- Ischemic stroke (IS)

- Genome-wide Association Study (GWAS)

- Next-Generation Sequencing (NGS)

- Rare variant association tests (RVAS)

- Whole exome sequencing (WES)

- Cardioembolism (CE)

- Large-artery atherosclerosis (LAA)

- Other determined (OD)

- Genome Analysis Toolkit (GATK)

- QualbyDepth (QD)

- Quality (QUAL)

- StrandOddsRatio (SOR)

- FisherStrand (FS)

- RMSMappingQuality (MQ)

- MappingQualityRankSumTest (MQRankSum)

- ReadPosRankSumTest (ReadPosRankSum)

- Principal component analysis (PCA)

- Glycosylphosphatidylinositol (GPI)

